# Geometric Deep Learning Methods for Improved Generalizability in Medical Computer Vision: Hyperbolic Convolutional Neural Networks in Multi-Modality Neuroimaging

**DOI:** 10.1101/2024.10.12.24315391

**Authors:** Cyrus Ayubcha, Sulaiman Sajed, Chady Omara, Anna B. Veldman, Shashi Bhushan Singh, Yashas Ullas Lokesha, Alex Liu, Mohammad Ali Aziz-Sultan, Timothy R. Smith, Andrew Beam

**Author notes:** Corresponding Author: Andrew Beam Harvard T.H. Chan School of Public Health, 677 Huntington Ave, Boston, MA 02115.

## Abstract

**Objective:** This study investigates the potential advantages of hyperbolic convolutional neural networks (HCNNs) over traditional convolutional neural networks (CNNs) in neuroimaging tasks.

**Materials and Methods:** We conducted a comparative analysis of HCNNs and CNNs across various medical imaging modalities and diseases, with a focus on a compiled multi-modality neuroimaging dataset. The models were assessed for performance parity, robustness to adversarial attacks, semantic organization of embedding spaces, and generalizability. Zero-shot evaluations were also performed with ischemic stroke non-contrast CT images.

**Results:** HCNNs matched CNN performance on less complex settings and demonstrated superior semantic organization, and robustness to adversarial attacks. While HCNNs equaled CNNs in out-of-sample datasets identifying Alzheimer’s disease, in zero-shot evaluations, HCNNs outperformed CNNs and radiologists.

**Discussion:** HCNNs deliver enhanced robustness and organization in the neuroimaging data. This likely underlies why while HCNNs perform similarly to CNNs with respect to in-sample tasks, they confer improved generalizability. Nevertheless, HCNNs encounter efficiency and performance challenges with larger, complex datasets. These limitations underline the need for further optimization of HCNN architectures.

**Conclusion:** HCNNs present promising improvements in generalizability and resilience for medical imaging applications, particularly in neuroimaging. Despite challenges with larger datasets, HCNNs enhance performance under adversarial conditions and offer better semantic organization, suggesting valuable potential in generalizable deep learning models in medical imaging and neuroimaging diagnostics.

## Background and Significance

Advances in computational power have facilitated the expansion of predictive deep learning models in many fields. While deep learning models are idealized as universal approximators, not all tasks are not equally appropriate for all neural network architectures; such that, misalignment may result in subpar empirical task performance.^1^ One notable instance of this may involve data structures with inherent hierarchical structure.^2^ Tree-like or hierarchical data structures have been shown to be represented with superior fidelity with less distortion in hyperbolic space as compared to Euclidian space.^3^ Specifically, the hyperbolic manifold allows for exponential scaling from the radial axis which mirrors the distance relationships found in hierarchical structures and accordingly, prevents distortion and information loss.^4^

There has been notable effort to utilize the superior data representation observed in hyperbolic spaces with deep learning algorithms. Over the last few years, many have attempted to operationalize constructs of hyperbolic space in a computationally efficient manner with the goal of reconstructing the fundamental functionality of neural network operations consistent with hyperbolic space.^2^ Thus, Hyperbolic Neural Networks (HNNs) were developed as an alternative to standard Euclidean neural networks.^5^ While most papers explore densely connected HNNs, there have been efforts to use alternative neural network architectures. For instance, Khrulkov et al developed a hybrid HNN that was applied to image datasets by utilizing a traditional Euclidean convolution structure and connected layer prior to mapping the resulting embeddings into hyperbolic space and conducting a multi-class logistic regression.^6^ Khrulkov et al showed superior performance to analogous Euclidean models in certain datasets (Caltech-UCSD Birds, DukeMTMC-reID dataset) as well as in few-shots classification. ^6^ They also showed that HNNs provided more intuitive organization of the classes in the embedding space likely explaining their improved out-of-sample and out-of-distribution performance.^6^

Given issues with numerical stability during training, most literature has moved to make HNNs more numerically stable.^7,8,9^ Guo et al proposed a clipping mechanism that would bound the embedding space so coerce numerically stabilize during training.^9^ These clipped HNNs were found to outperform standard HNNs on various benchmarks, including CIFAR10, CIFAR100, and ImageNet, and demonstrated better adversarial robustness and out-of-distribution detection.^9^ Unlike unclipped HNNs, clipped HNNs achieved performance on par with ENNs in data settings without natural tree structures. The use of the Lorentz model of hyperbolics spaces, which has different mathematical constructions for the computational operations of the neural network as compared to the Poincaré ball model, has also been proven to reduce numerical instability.^8,10^ Finally, there have also been efforts to translate common Euclidian convolutional neural network operations into hyperbolic space where the fully hyperbolic convolutional neural networks (HCNN) could be contained to hyperbolic space in an end-to-end fashion.^11^ This prevents the need for mapping between Euclidean and hyperbolic spaces so to limit numerical instability.

Nevertheless, the literature remains uncertain in the fully exploring the value of HNNs. The original seminal paper in hyperbolic imaging embeddings suggests that most imaging datasets have some degree of implicit hierarchical structure.^6^ Other studies found poor performance in settings without any natural hierarchical structures as compared to Euclidean counterparts until Guo et al suggested that clipped HNNs achieved similar performance in certain settings.^9^ In totality, there is a clear need to further robust evaluation of HCNNs in various domains to weight the possible benefits and limitation of these evolving models.

One field of application includes medical imaging where the successful application of computer vision algorithms has been keenly appreciated.^12^ To our knowledge, only one study has been performed utilizing HNNs for classification of medical imaging data. Utilizing Khrulkov et. al’s hybrid paradigm, Yu et al introduces a hyperbolic prototype network capable of jointly learning image embeddings and class prototypes in a shared hyperbolic space, guided by an error construction mechanism derived from a prior known class hierarchy.^13^ Their approach preserved the semantic class relationships of dermatoscope images in the hyperbolic embedding space and found superior performance in classification as compared to analogous Euclidean approaches, though with lower space curvature hyperparameters.^13^

Given the success of present day convolutional neural networks in imaging recognition tasks, there has been a greater interest in developing models with both wide spanning capabilities across a variety of tasks or classes; and those durable to a variety of clinical settings where models may encounter rare patient morphologies or imperfect images.^14–17^ We explore the value of HCNNs by evaluating the performance of clipped HCNNs relative to their Euclidean counterparts in neuroimaging settings as well as other multi-modality medical imaging databases. Agnostic to any prior data hierarchy, we conduct an evaluation of in sample performance across three medical imaging datasets of varying complexity. We evaluate the neuroimaging models not only in their respective classification performance but also in their organization of embedding spaces, durability to adversarial attacks, and out-of-sample as well as zero-shot generalizability.

## Materials and Methods

### Data Sources

We develop a core neuro-imaging dataset to evaluate the HCNN and CNN models: Multi-Modality Neuroimaging (MMN) Dataset composed of 72,634 images of 42 total classes. The images were acquired from previously open-source databases which included various neurological diseases including ischemic stroke^18^, hemorrhagic stroke^19^, metastasis^20^, tumor^21^, schizophrenia (COBRE, MCICShare)^22^, and Alzheimer’s disease (AD) ^23^ across computed tomography (CT) and Magnetic Resonance Imaging (MRI) modalities. Images not from a peer-reviewed source, were independently confirmed to be correctly classified according to a qualified radiologist.

Certain classes in the neuroimaging dataset were manually composed due to either overlapping classes or, occasionally, to better capture disease signals. Specifically, the “normal” categories are composed from multiple appropriate non-diseased classes in the datasets above. For instance, patients in the schizophrenia databases defined as “normal” patients was moved into the respective normal classes once we excluded the possibility of them joining another diseased class in the database. Schizophrenia positive scans were restricted to four of the median axial scans for each patients to better capture morphological abnormalities commonly conferred by patients with schizophrenia.^31^

We also constructed two additional datasets of other medical imaging types to conduct further analyses in larger and smaller class settings: Miniature Multi-Disease Dataset (MMD), and Multi-Disease Dataset (MD). The MD is composed of 89,496 images of 78 total classes. These images were acquired from previously published open-source databases which include Chest X-Rays^24^, Fundoscopy^25^, Gastrointestinal Scopes^26^, Musculoskeletal X-Rays^27^, Neuroimaging and Dermatoscopy.^28–30^ Finally, the MMD was restricted to a smaller subset of the MMD with a total of 19,880 unique images across 16 total classes. The classes and their respective imaging data can be observed in **Table 1**.

**Table 1:** Dataset Characteristics.

| Miniature Multi-Disease: Classes | Images | Multi-Modality Neuroimaging: Classes | Images | Multi-Disease Dataset: Classes | Images |
| --- | --- | --- | --- | --- | --- |
| Derm: Actinic Keratosis | 867 | AD Moderate MRI T1 | 896 | Derm: Actinic Keratosis | 867 |
| Derm: Basal Cell Carcinoma | 3323 | AD Severe MRI | 64 | Pulm: Bacterial Pneumonia | 2780 |
| Derm: Benign Keratosis | 2624 | AD Mild MRI T1 | 2240 | Derm: Basal Cell Carcinoma | 3323 |
| Derm: Dermatofibroma | 239 | Hemorrhagic Stroke Epidural CT Bone | 167 | Derm: Benign Keratosis | 2624 |
| Derm: Melanoma | 4522 | Hemorrhagic Stroke Intraparenchymal CT Bone | 52 | Derm: Dermatofibroma | 239 |
| Derm: Melanocytic Nevi | 12875 | Hemorrhagic Stroke Intraventricular CT Bone | 13 | Neuro: Epidural Hemmorhagic Stroke CT Bone | 167 |
| Derm: Squamous Cell Carcinoma | 628 | Hemorrhagic Stroke Subarachnoid CT Bone | 9 | Neuro: Intraparenchymal Hemmorhagic Stroke CT Bone | 52 |
| Derm: Vascular Lesion | 253 | Hemorrhagic Stroke Subdural CT Bone | 52 | Neuro: Subdural Hemmorhagic Stroke CT Bone | 52 |
| Gastro: Dyed Lifted Polyps | 1000 | Hemorrhagic Stroke Epidural CT Brain | 167 | Neuro: Epidural Hemmorhagic Stroke CT Brain | 167 |
| Gastro: Dyed Resection Margins | 1000 | Hemorrhagic Stroke Intraparenchymal CT Brain | 52 | Neuro: Intraparenchymal Hemmorhagic Stroke CT Brain | 52 |
| Gasto: Esophagitis | 1000 | Hemorrhagic Stroke Intraventricular CT Brain | 13 | Neuro: Subdural Hemmorhagic Stroke CT Brain | 52 |
| Optho: Normal Fundus | 2873 | Hemorrhagic Stroke Subarachnoid CT Brain | 9 | Neuro: Ischemic Stroke DWI | 1012 |
| Gastro: Normal Cecum | 1000 | Hemorrhagic Stroke Subdural CT Brain | 52 | Neuro: Ischemic Stroke Flair | 1002 |
| Gastro: Normal Pylorus | 1000 | Ischemic Stroke MRI DWI | 1012 | Derm: Melanoma | 4522 |
| Gasto: Normal Z-Line | 1000 | Ischemic Stroke MRI Flair | 1002 | Neuro: Metastasis Flair | 4248 |
| Gastro: Polyps | 1000 | Metastasis MRI Flair | 4248 | Neuro: Metastasis T1 | 4248 |
|  |  | Metastasis MRI T1C | 4248 | Neuro: Metastasis T1C+ | 4248 |
|  |  | Metastasis MRI T1 | 4248 | Pulm: Normal CXR | 1583 |
|  |  | Normal CT Bone | 1495 | Derm: Melanocytic Nevi | 12875 |
|  |  | Normal CT Brain | 1494 | Neuro: Normal CT Bone | 1495 |
|  |  | Normal MRI DWI | 1406 | Neuro: Normal CT Brain | 1494 |
|  |  | Normal MRI Flair | 14949 | Neuro: Normal DWI | 1406 |
|  |  | Normal MRI T1 | 17925 | Neuro: Normal_Flair | 4949 |
|  |  | Normal MRI T1C+ | 13941 | Neuro: Normal T1 | 7925 |
|  |  | Normal MRI T2 | 18 | Neuro: Normal T1C+ | 3941 |
|  |  | Schizophrenia MRI DWI | 471 | Neuro: Normal T2 | 18 |
|  |  | Schizophrenia MRI T1 | 1314 | Derm: Squamous Cell Carcinoma | 628 |
|  |  | Glioma MRI T1C+ | 152 | Neuro: Schizophrenia DWI | 471 |
|  |  | Meningioma MRI T1C+ | 233 | Neuro: Schizophrenia T1 | 1314 |
|  |  | Neurocitoma MRI T1C+ | 76 | Neuro: Glioma T1 | 152 |
|  |  | Other Lesions MRI T1C+ | 9 | Neuro: Meningioma T1 | 233 |
|  |  | Schwannoma MRI T1C+ | 36 | Neuro: Neurocitoma T1 | 76 |
|  |  | Glioma MRI T1 | 65 | Neuro: Schwannoma T1 | 36 |
|  |  | Meningioma MRI T1 | 141 | Neuro: Glioma T1C+ | 65 |
|  |  | Neurocitoma MRI T1 | 39 | Neuro: Meningioma T1C+ | 141 |
|  |  | Other Lesions MRI T1 | 27 | Neuro: Neurocitoma T1C+ | 39 |
|  |  | Schwannoma MRI T1 | 31 | Neuro: Schwannoma T1C+ | 31 |
|  |  | Glioma MRI T2 | 9 | Neuro: Glioma T2 | 67 |
|  |  | Meningioma MRI T2 | 67 | Neuro: Meningioma T2 | 145 |
|  |  | Neurocitoma MRI T2 | 145 | Neuro: Schwannoma T2 | 33 |
|  |  | Other Lesions MRI T2 | 14 | Derm: Vascular Lesion | 253 |
|  |  | Schwannoma MRI T2 | 33 | Pulm: Viral Pneumonia | 1493 |
|  |  |  |  | Optho: Branch Retinal Vein Occlusion | 19 |
|  |  |  |  | Optho: Cataract | 287 |
|  |  |  |  | Optho: Diabetic Retinopathy | 53 |
|  |  |  |  | Optho: Drusen | 148 |
|  |  |  |  | Optho: Dry Age-Related Macular Degeneration | 202 |
|  |  |  |  | Gastro: Dyed Lifted Polyps | 1000 |
|  |  |  |  | Gastro: Dyed Resection Margins | 1000 |
|  |  |  |  | Optho: Epiretinal Membrane | 140 |
|  |  |  |  | Gasto: Esophagitis | 1000 |
|  |  |  |  | Optho: Glaucoma | 213 |
|  |  |  |  | MSK: Hand Normal | 877 |
|  |  |  |  | MSK: Hand Fractured | 379 |
|  |  |  |  | MSK: Hand Shoulder Normal | 180 |
|  |  |  |  | MSK: Hand Shoulder Fractured | 53 |
|  |  |  |  | MSK: Hip Normal | 169 |
|  |  |  |  | MSK: Hip Fractured | 13 |
|  |  |  |  | Optho: Hypertensive Retinopathy | 123 |
|  |  |  |  | Optho: Macular Epiretinal Membrane | 140 |
|  |  |  |  | Optho: Maculopathy | 23 |
|  |  |  |  | Optho: Mild Nonproliferative Retinopathy | 464 |
|  |  |  |  | Optho: Moderate Non-Proliferative Retinopathy | 798 |
|  |  |  |  | Optho: Myelinated Nerve Fibers | 68 |
|  |  |  |  | Optho: Normal Fundus | 2873 |
|  |  |  |  | Gastro: Normal Cecum | 1000 |
|  |  |  |  | Gastro: Normal Pylorus | 1000 |
|  |  |  |  | Gasto: Normal Z-Line | 1000 |
|  |  |  |  | Optho: Pathological Myopia | 231 |
|  |  |  |  | Gastro: Polyps | 1000 |
|  |  |  |  | Optho: Refractive Media Opacity | 54 |
|  |  |  |  | Optho: Severe Nonproliferative Retinopathy | 144 |
|  |  |  |  | Gastro: Ulcerative Colitis | 1000 |
|  |  |  |  | Optho: Vitreous Degeneration | 58 |
|  |  |  |  | Optho: Wet Age-Related Macular Degeneration | 41 |

Out-of-sample data were acquired from the T-1 MRI sequences of the OASIS-1 cross-sectional cohort imaging study^32^. We derived one mid-brain axial slice from each patient in the study and defined an AD positive case as an individual with a Mini-Mental State Examination (MMSE) score of below 25 and negative otherwise. In total, this dataset entailed a total of 436 subjects where 40 subjects were AD positive and 396 were AD negative. The zero-shot dataset was acquired from the Mass General Brigham (MGB) Research Patient Data Registry (RPDR) with MGB Institutional Review Board approval. We retrospectively selected individuals from January 2004 to December 2022 who received Non-Contrast Computed Tomography (NCCT) brain scan upon presenting to the emergency department. We further restricted ourselves to those diagnosed with acute ischemic strokes on Magnetic Resonance Imaging (MRI) brain scans within 7 days of their original presentation to the emergency department. We include the full axial scans of the 151 patients for a total of 23,371 images. We also document the radiologist reports and their ability to correctly able to diagnose the ischemic stroke on NCCT. We label the axial images as positive if they included regions implicated by the positive MRI reads for that patient where all other images are deemed negative for that patient.

### Processing and Models

All training data underwent uniform pre-processing transformations prior to their use in the model including normalization, rotation, and horizontal flip. Furthermore, to homogenize the diverse presentation of brain imaging data, we conducted skull-stripping when appropriate. For all the models in this study, we utilize a Res-Net-18 (RN18) backbone where we define the baseline Euclidean CNNs as the RN18. The HCNN models are constructed as hybrid models with an identical convolution structure as the CNN. However, the HCNN model acquires the subsequent embeddings and translates them into the Lorentz space with an exponent mapping procedure where the remaining dense layers of the RN18 backbone reside. Other features of our hybrid HCNN construct include trainable curvature, feature-clipping^9^, and Euclidean reparameterizations^10^ which are documented by the Bdeir et al^33^ code base we utilize for our study.

### Evaluation

To examine the performance of each model in the respective dataset, we reproduce cross-entropy loss, top-1 accuracy, and top-5 accuracy. Utilizing the average embedding position of each class we construct a low dimensional representation using T-SNE algorithms with the Euclidean and Lorentz models, respectively. We also utilize hierarchical clustering procedures with the average class embedding positions for each model which allow us to derive a dendrogram of the inter-class relationships learned by the respective models.

To examine the interpretability of the embedding space, we compare the geodesic distance matrices of average class embeddings from the respective Euclidean and Euclidean-Lorentz models to a ground truth hierarchical distance matrix. We derive the ground truth distance matrix based on the sets of categorical variables as descriptors of the imaging category and disease type (refer to Appendix). We first normalize all distance matrices and compute the absolute pairwise difference between the model distance matrix and the ground truth distance matrix. We also derive the Spearman’s rank correlation coefficient between the respective model and ground truth distances matrices based on the ranked distance of the pairwise classes.

To evaluate out-of-sample accuracy, we utilize a single median axial slice in each OASIS-1 subject and consider a positive diagnosis as one that ascribes an AD-related class to the scan and a negative diagnosis as a normal T1 MRI predicted class. Next, we evaluate zero-shot accuracy by defining a true positive diagnosis read as having an axial NCCT image predicted as an ischemic or hemorrhagic stroke class by the model where the ground truth was positive. At a patient level, we identify accuracy as a patient having at least one true positive. We define a true negative as a normal CT class. Then we identify overall image-based accuracy as the number of true positives and true negatives over the overall number of images. Finally, we conduct a Projected Gradient Descent (PGD) adversarial attack to assess the comparative durability of each model to more extreme distortions in data. Note that all 95% confidence intervals are derived from a respective bootstrapping procedure (n=1000).

## Results

The performance of the Euclidean model was generally matched by the Euclidean-Lorentz model in the MMN and the MMD datasets with minimal differences in Top-1 accuracy and identical Top-5 accuracy (**Table 2**). More broadly, however, the Euclidean model generally began to outperform the Euclidean-Lorentz model as the number of images and class size of the dataset increased; this is most prominent in the MD dataset (**Figure 1**). Interpreting the low-dimensional T-SNE representation of the average embedding class as well as the respective dendrogram, the Euclidean-Lorentz model appears to have a more reasonable distribution of the embedding space based on prior understanding of the inter-relationships between the imaging classes in the MMN (**Figures 2 and 3**).

**Table 2:** Model Characteristics and Performance.

| Dataset | Class Size | Sample Size | Model Type | Trainable Parameters | Cross Entropy Loss | Top 1 Accuracy | Top 1 Accuracy: 95% Confidence Intervals | Top 5 Accuracy |
| --- | --- | --- | --- | --- | --- | --- | --- | --- |
| MMN | 42 | 72,634 | Euclidean ResNet 18 | 11,189,226 | **0.05** | **97.4** | **97.3 - 97.5** | **99.9** |
| MMN | 42 | 72,634 | Euclidean-Lorentz ResNet 18 | 11,189,227 | 0.12 | 96.2 | 96.1 - 96.4 | **99.9** |
| MD | 75 | 89,496 | Euclidean ResNet 18 | 11,207,307 | **0.26** | **90.8** | **90.6 - 91.0** | **99.6** |
| MD | 75 | 89,496 | Euclidean-Lorentz ResNet 18 | 11,207,308 | 0.47 | 85.6 | 85.4 - 85.9 | 99.0 |
| MMD | 16 | 19,880 | Euclidean ResNet 18 | 11,177,040 | **0.11** | **97.3** | **97.1 - 97.4** | **99.9** |
| MMD | 16 | 19,880 | Euclidean-Lorentz ResNet 18 | 11,177,041 | **0.11** | 97.0 | **96.8 - 97.1** | **99.9** |

**Figure 1:**
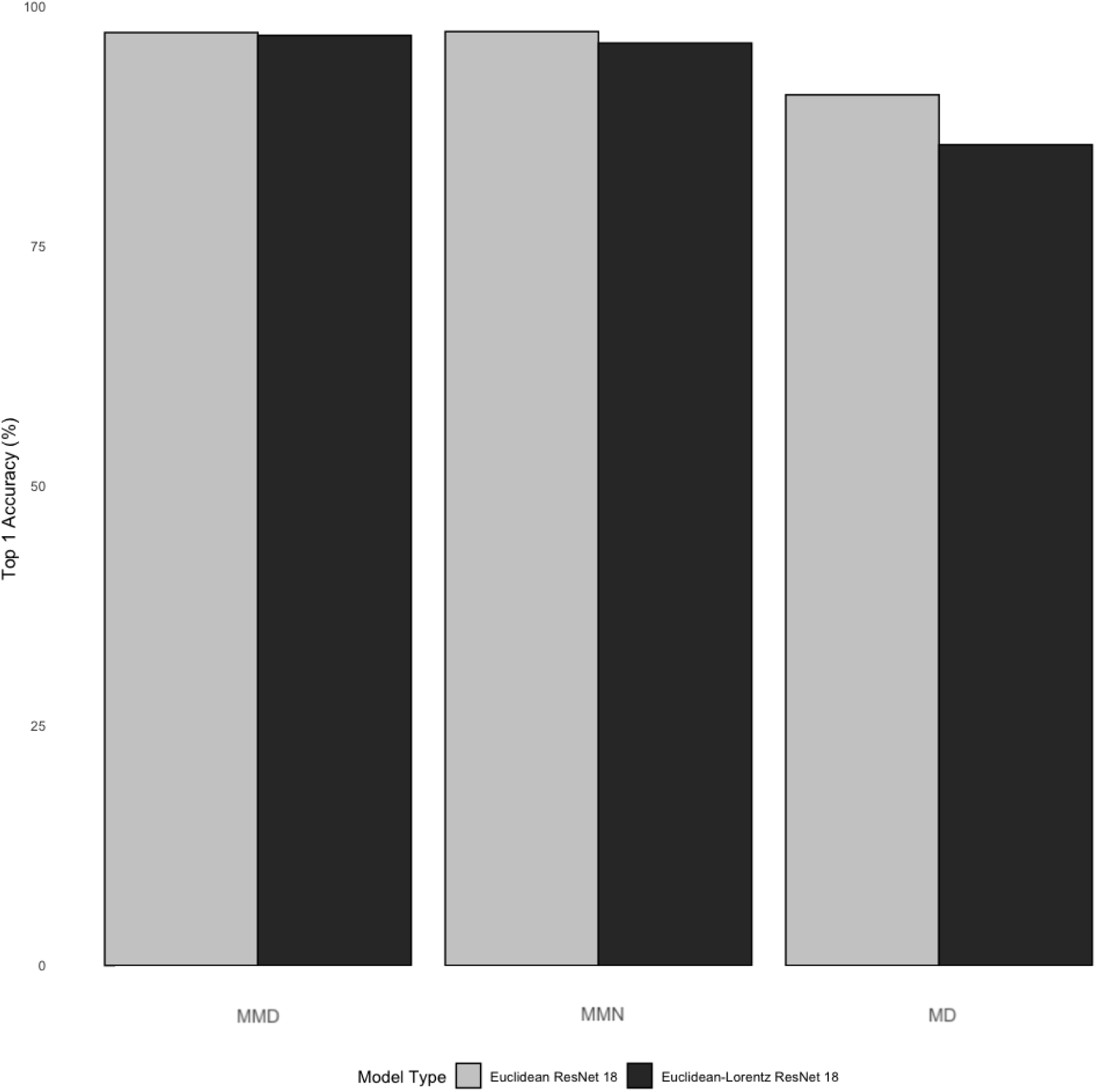
Relative Model Performance Across Datasets. The bar plot above shows the top-1 accuracy metrics with 95% confidence intervals for the Euclidean ResNet 18 and the Euclidean-Lorentz ResNet 18 across the three datasets increasing in size from left to right (i.e., Miniature Multi-Disease (MDD) Dataset, Multi-Modality Neuroimaging (MMN) Dataset, and Multi-Disease (MD) Dataset).

**Figure 2:**
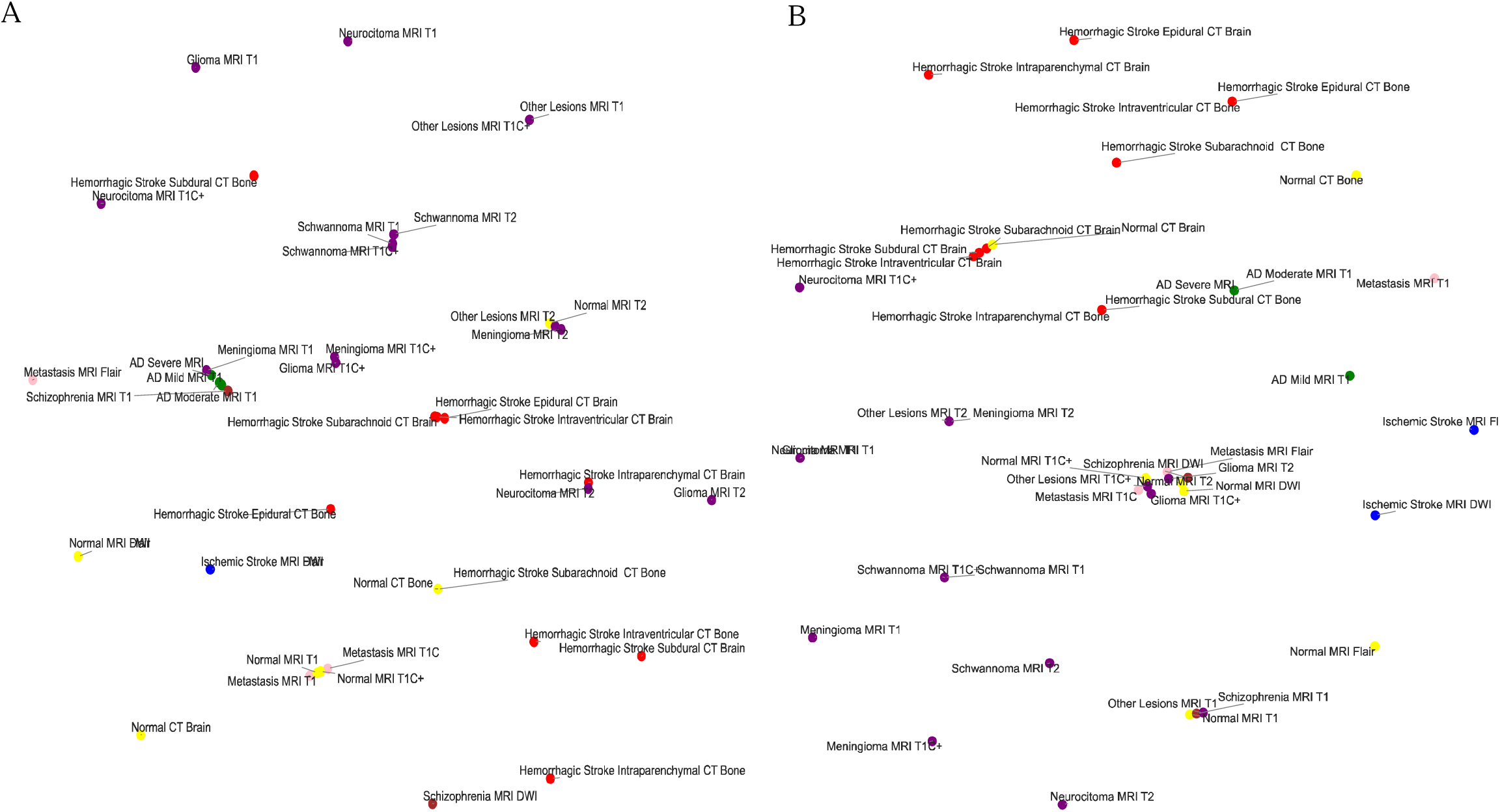
Euclidean and Hyperbolic Models T-SNE in the Neuroimaging Dataset. The figure shows the low dimensional representation T-SNE of the average class embedding space from the Euclidean ResNet 18 (A) and the Euclidean-Lorentz ResNet 18 (B) in the Multi-Modality Neuroimaging (MMN) Dataset

**Figure 3:**
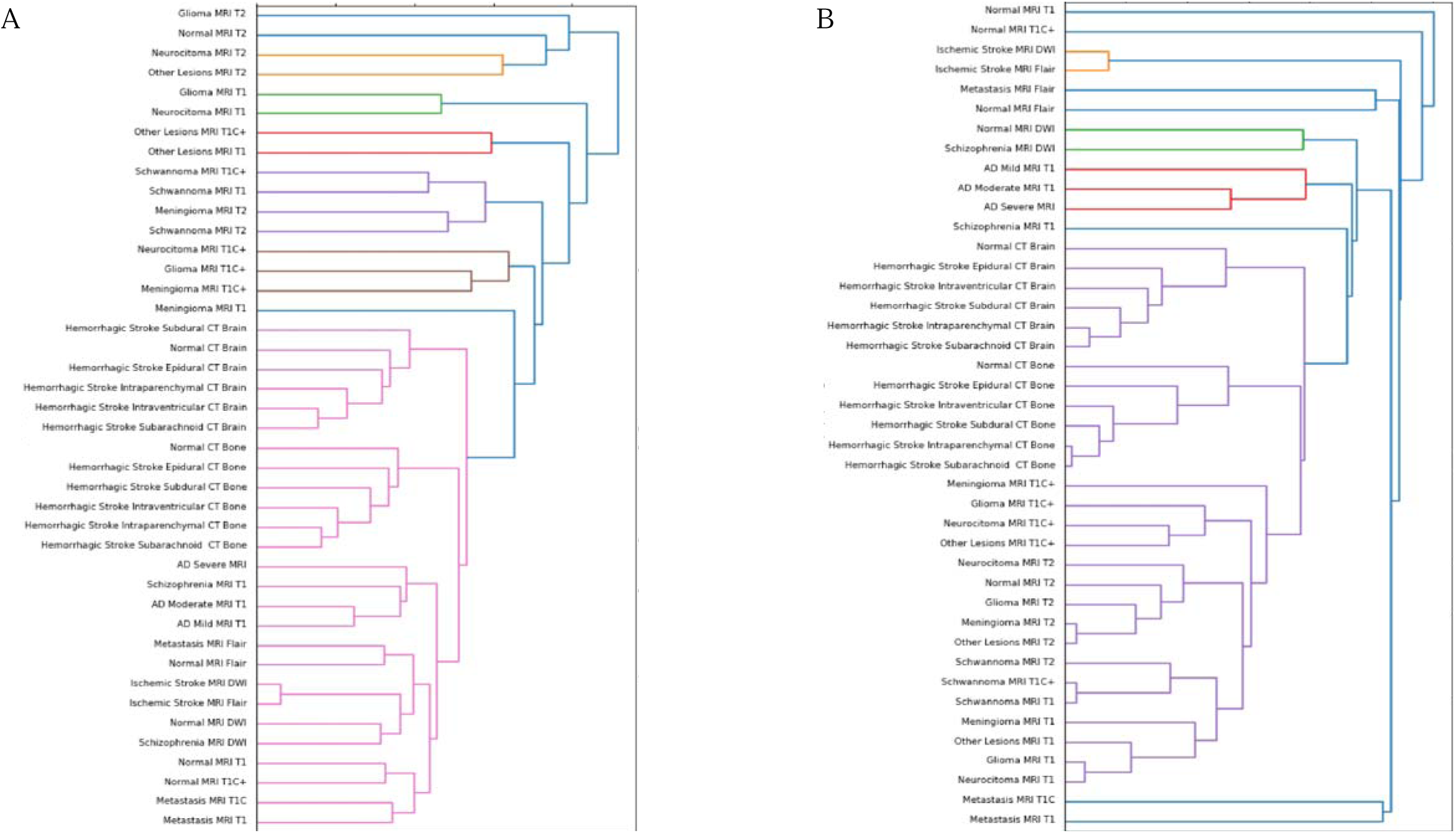
Euclidean and Hyperbolic Models Dendrograms in the Neuroimaging Dataset. The figure illustrates the hierarchical clustering dendrogram of the average class embedding space of the Euclidean ResNet 18 (A) and the Euclidean-Lorentz ResNet 18 (B) in the Multi-Modality Neuroimaging (MMN) Dataset

When compared to the known ground truth distance matrix, the distinction between the two models becomes more apparent. The mean absolute difference between the respective Euclidean and Euclidean-Lorentz models compared to the ground truth distance matrix for the MMN dataset was highest in the Euclidean model (0.290 ± 0.005). In contrast, the mean absolute difference in the Euclidean-Lorentz model was significantly lower (0.158 ± 0.003), with a two-sample t-test p-value of < 0.0001. Spearman’s rank correlation findings show that the Euclidean model exhibited a weak correlation with the ground truth ranking (correlation coefficient = 0.021, p = 0.3783), while the Euclidean-Lorentz model showed a stronger correlation (correlation coefficient = 0.328, p < 0.0001). These results indicate that the Euclidean-Lorentz model not only has a lower mean absolute difference compared to the ground truth but also demonstrates a stronger correlation to the ground-truth class ranks.

Compared to the radiologist’s performance, which identified 82 of the 151 patients (0.53), the Euclidean model performed worse, identifying only 62 stroke patients (0.41), while the Euclidean-Lorentz model outperformed by identifying 94 (0.62). Across all images, the Euclidean-Lorentz model achieved a higher overall accuracy (0.50) than the Euclidean model (0.45) (**Figure 4**).

**Figure 4:**
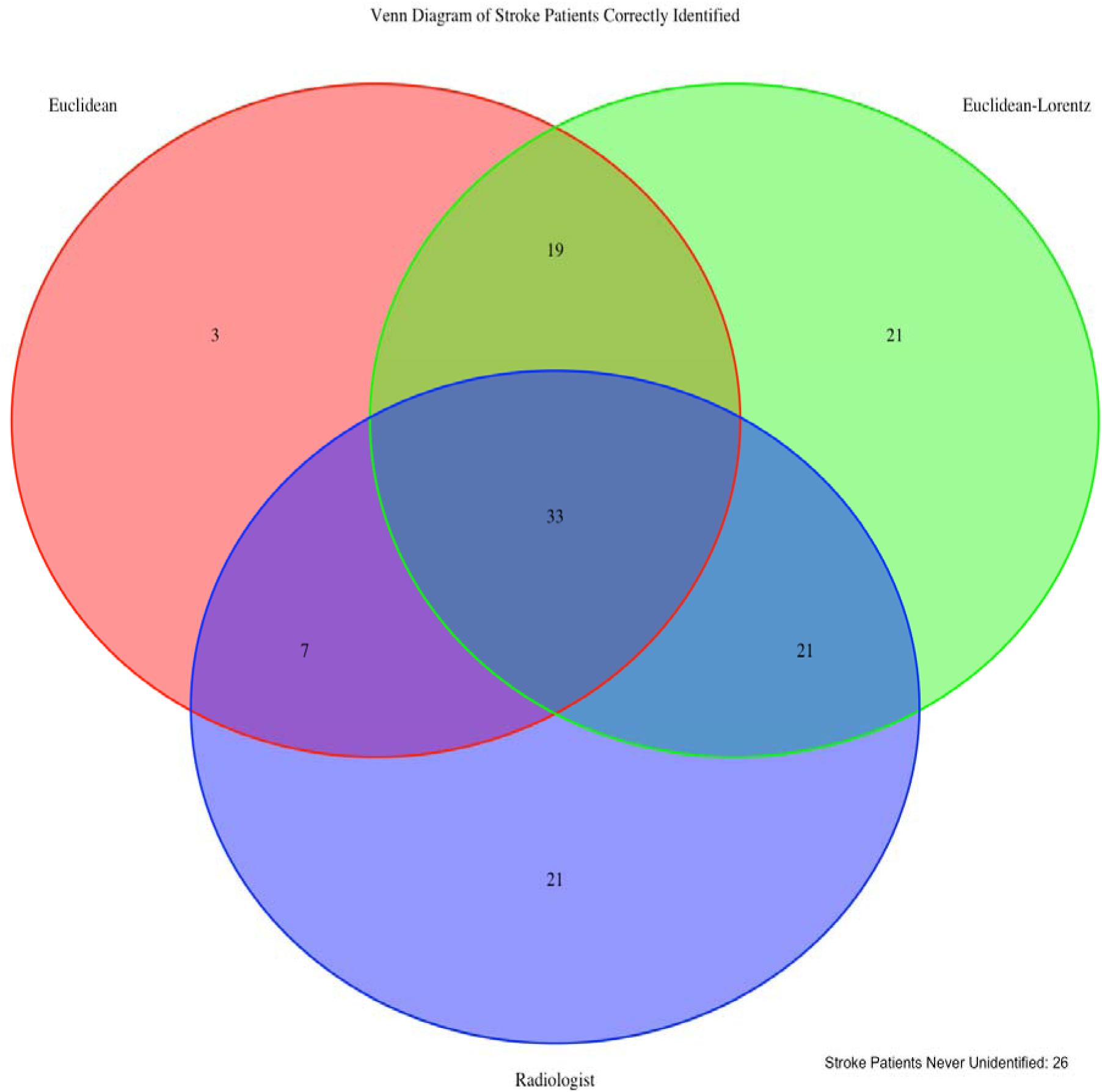
Zero-Shot Identification of Stroke Patients. The diagram above shows the how many of the zero-shot stroke patients were able to be identified across in the Euclidean and Euclidean-Lorentz models as well as by human radiologists with their emergent non-contrast brain CT imaging. We also note that 26 patients were not identified in any of the three approaches.

In the out-of-sample dataset, both models were able to correctly identify the modality of the axial images with a 100% identification rate. The Euclidean-Lorentz model and the Euclidean model achieved a Top-1 accuracy of 0.54 (95% CI: 0.44, 0.64) and 0.55 (95% CI: 0.45, 0.65), respectively, suggesting statistically indistinguishable performance in this out-of-sample dataset. Within the NCCT ischemic stroke dataset, the negative cases were technically a class already observed by the models, so we used this as an additional out-of-sample experiment where the models were tasked with correctly identifying negative NCCT slices as normal CT images. The Euclidean model achieved an accuracy among the negative axial NCCT images of 0.81 (95% CI: 0.81 - 0.82), which was statistically comparable to the Euclidean-Lorentz model, which reached an accuracy of 0.82 (95% CI: 0.82 - 0.83).

Interestingly, the PGD adversarial attack analysis suggests that the Euclidean-Lorentz model often outperforms its Euclidean counterpart in the larger MMN and MD datasets with respect to Top-1 and Top-5 accuracy metrics (**Table 3**). The performance becomes more similar across the two models in the smaller MMD dataset.

**Table 3:**
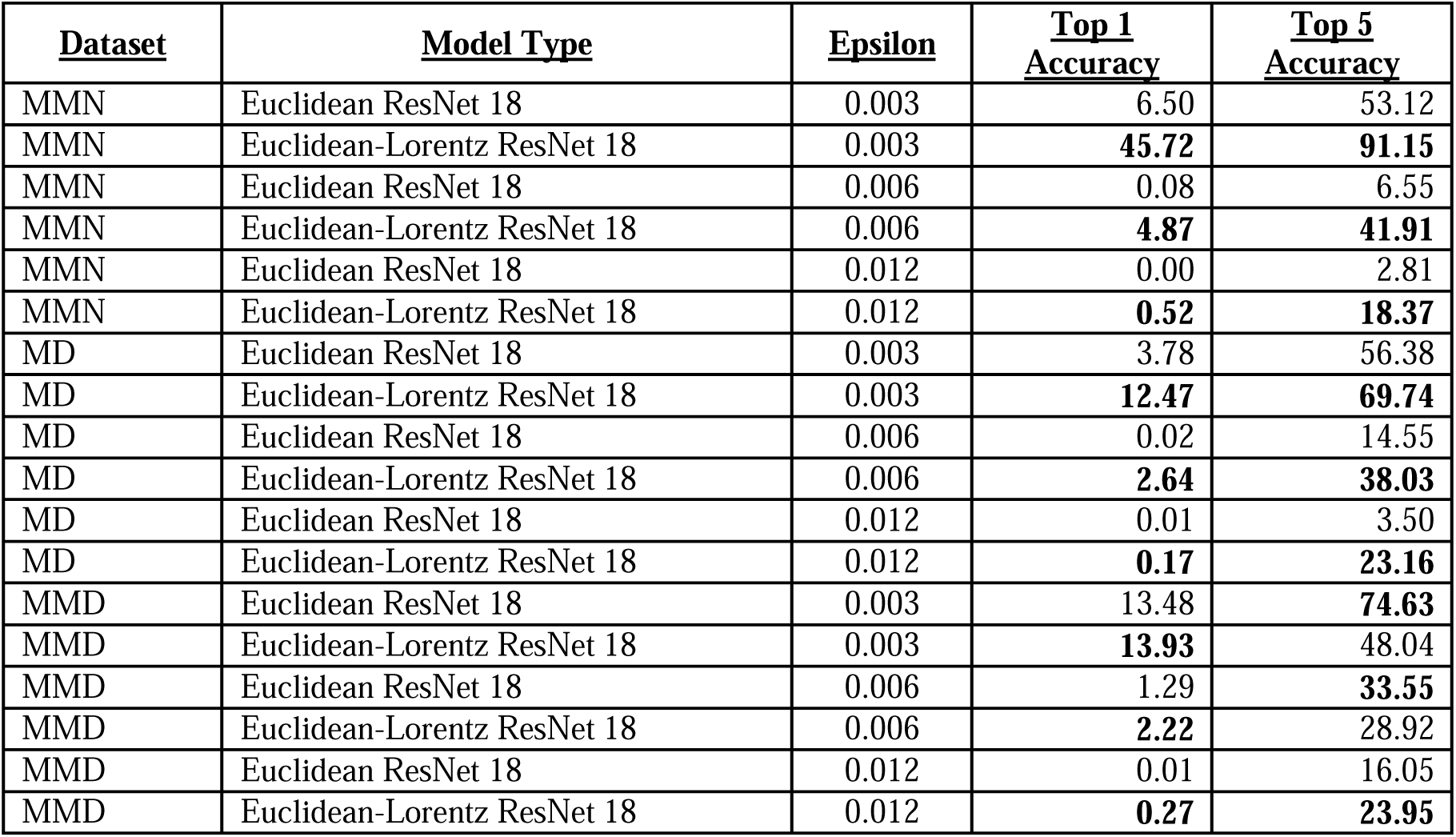
Projected Gradient Descent Adversarial Attack.

| <u>Dataset</u> | <u>Model Type</u> | <u>Epsilon</u> | <u>Top 1 Accuracy</u> | <u>Top 5 Accuracy</u> |
| --- | --- | --- | --- | --- |
| MMN | Euclidean ResNet 18 | 0.003 | 6.50 | 53.12 |
| MMN | Euclidean-Lorentz ResNet 18 | 0.003 | **45.72** | **91.15** |
| MMN | Euclidean ResNet 18 | 0.006 | 0.08 | 6.55 |
| MMN | Euclidean-Lorentz ResNet 18 | 0.006 | **4.87** | **41.91** |
| MMN | Euclidean ResNet 18 | 0.012 | 0.00 | 2.81 |
| MMN | Euclidean-Lorentz ResNet 18 | 0.012 | **0.52** | **18.37** |
| MD | Euclidean ResNet 18 | 0.003 | 3.78 | 56.38 |
| MD | Euclidean-Lorentz ResNet 18 | 0.003 | **12.47** | **69.74** |
| MD | Euclidean ResNet 18 | 0.006 | 0.02 | 14.55 |
| MD | Euclidean-Lorentz ResNet 18 | 0.006 | **2.64** | **38.03** |
| MD | Euclidean ResNet 18 | 0.012 | 0.01 | 3.50 |
| MD | Euclidean-Lorentz ResNet 18 | 0.012 | **0.17** | **23.16** |
| MMD | Euclidean ResNet 18 | 0.003 | 13.48 | **74.63** |
| MMD | Euclidean-Lorentz ResNet 18 | 0.003 | **13.93** | 48.04 |
| MMD | Euclidean ResNet 18 | 0.006 | 1.29 | **33.55** |
| MMD | Euclidean-Lorentz ResNet 18 | 0.006 | **2.22** | 28.92 |
| MMD | Euclidean ResNet 18 | 0.012 | 0.01 | 16.05 |
| MMD | Euclidean-Lorentz ResNet 18 | 0.012 | **0.27** | **23.95** |

## Discussion

Our empirical analysis study elucidates several important insights into the comparative performance of clipped Euclidean-Lorentz HCNNs and Euclidean CNNs in neuroimaging tasks as well as other medical imaging settings. The results suggest parity in performance between the two neural network approaches in smaller, less complex datasets. We further note distinct semantic organization within the respective embedding spaces, where the HCNN aligned better with ground truth relations between the neuroimaging classes. In assessing generalizability, the HCNN achieved similar out-of-sample performance in identifying AD and normal NCCT images but greatly improved zero-shot performance in identifying ischemic stroke on NCCT.

The cross-entropy loss and top-1 accuracy metrics followed a similar trend across the three medical imaging datasets. Notably, these metrics were identical or similar in datasets where the CNN achieved higher performance (>95% accuracy). However, as the complexity and size of the datasets grew larger, there was a precipitous drop in HCNN performance compared to the CNN. Interestingly, despite the difference in loss in the MMN dataset, the top-1 accuracy between the two models was more similar, unlike in the MD dataset. In both settings of performance parity and disparity, the top-5 accuracy metric across the two models was nearly identical in all three datasets, perhaps due to the improved generalizability of HCNNs, which we explore further.

Achieving parity replicates the findings from Guo et al., which demonstrate that clipped HCNNs achieve similar performance in data settings without strong hierarchy.^9^ Nevertheless, we illustrate that the performance of the HCNN suffers compared to the CNN when applied to larger datasets with seemingly more difficult tasks. Given the similarity in model size across the three datasets, our findings may suggest that the HCNNs, as currently constructed, are less efficient with their trainable parameters, contrary to prior literature.^5^

One of the known features of HCNNs is the improved preservation of hierarchical data structures, as reflected by the organized embedding space.^2,6^ Low-dimensional T-SNE representations of the embedding space suggest a stratification of classes in the MMN dataset, often by modality first and then disease type, in both models. Similarities in class grouping may be starker in the Euclidean-Lorentz model, as observed in the hierarchical clustering dendrogram from the embedding space.

Nevertheless, the noted limitations of low-dimensional representations may offer a distorted view of the true geodesic distances between the average class embeddings.^34^ To explore whether the two models developed meaningfully distinct organizations of embedding space in a more robust fashion, we derived a respective geodesic distance matrix for the average class embedding in both MMN models. We then compared the pairwise distance matrix from each model against a constructed ground truth difference matrix between the classes. Using the pairwise distance differences as well as the Spearman’s rank correlation coefficient, we show that the HCNN, and not the CNN, better aligned with our known semantic understanding of the class relationships.

While we observe superior learning and conservation of known class structures in neuroimaging data, we further explored the tangible value of this distinguishing feature. One of the more important aspects of any diagnostic medical imaging algorithm is its ability to function with out-of-sample and out-of-distribution imaging data.^35^ We specifically find that the MMN HCNN performed similarly to the CNN in the OASIS I and stroke-negative NCCT datasets, despite poorer HCNN performance in top-1 accuracy and loss in the MMN dataset.

We also find that in zero-shot performance, the HCNN unequivocally outperforms not only the CNN but also the trained radiologists by a significant margin. Finally, the HCNN has shown consistently increased durability to adversarial attacks, which may be relevant when confronted with imaging artifacts, image quality disparities between scanners, or image corruption that may or may not be perceptible.^36,37^ As suggested by prior studies in non-medical imaging settings^6,8,9^, we show that the neuroimaging HCNN has improved generalizability with respect to out-of-sample, zero-shot, and adversarial attack performance.

Our study should be interpreted with certain limitations. While recent studies have attempted to move convolutional functions into hyperbolic space,^11,33^ we observed significant numerical instability with these methods. Moreover, the computational efficiency of HCNNs was dramatically lower, with convergence requiring three to four times more epochs with similar hyperparameters on larger datasets, limiting our ability to use datasets larger than those in this study. We tested task complexity and size concurrently across the three datasets, but future work should explore scalability and complexity further. Additionally, we are restricted to speculating on the performance of clipped hybrid HCNNs in the context of our included medical imaging classes and can only speak to generalizability in terms of the out-of-sample and zero-shot datasets used.

## Conclusion

In this study, we show that agnostic HCNN models demonstrated superior ability to learn and retain the native hierarchical structure in the neuroimaging dataset compared to Euclidean CNNs. Importantly, the HCNN achieved disproportionately superior performance in adversarial attack experiments and zero-shot settings, outperforming both radiologists and CNNs. The neuroimaging HCNN also achieved parity in in-sample performance with out-of-sample data but showed depreciating performance in more complex medical imaging task settings with larger datasets. These findings suggest that improvements in the efficiency and scalability of HCNNs are needed to achieve parity with CNNs. However, HCNNs provided notable value in their generalizability in medical imaging settings with multi-modal and multi-disease neuroimaging data.

## Data Availability

Compiled data from publicly available sources is available upon request.

## Appendix I. Neuroimaging Class Table Characteristics Tables

I. Neuroimaging Class Table Characteristics Tables
| Multi-Modality Neuroimaging: Classes | Modality General | Modality Specific | Disease State | Disease Specific |
| --- | --- | --- | --- | --- |
| AD Moderate MRI T1 | mri | t1 | diffuse | ad |
| AD Severe MRI | mri | t1 | diffuse | ad |
| AD Mild MRI T1 | mri | t1 | diffuse | ad |
| Hemorrhagic Stroke Epidural CT Bone | ct | ctbo | lesion | hemm |
| Hemorrhagic Stroke Intraparenchymal CT Bone | ct | ctbo | lesion | hemm |
| Hemorrhagic Stroke Intraventricular CT Bone | ct | ctbo | lesion | hemm |
| Hemorrhagic Stroke Subarachnoid CT Bone | ct | ctbo | lesion | hemm |
| Hemorrhagic Stroke Subdural CT Bone | ct | ctbo | lesion | hemm |
| Hemorrhagic Stroke Epidural CT Brain | ct | ctbr | lesion | hemm |
| Hemorrhagic Stroke Intraparenchymal CT Brain | ct | ctbr | lesion | hemm |
| Hemorrhagic Stroke Intraventricular CT Brain | ct | ctbr | lesion | hemm |
| Hemorrhagic Stroke Subarachnoid CT Brain | ct | ctbr | lesion | hemm |
| Hemorrhagic Stroke Subdural CT Brain | ct | ctbr | lesion | hemm |
| Ischemic Stroke MRI DWI | mri | dwi | lesion | isch |
| Ischemic Stroke MRI Flair | mri | flair | lesion | isch |
| Metastasis MRI Flair | mri | flair | lesion | met |
| Metastasis MRI T1C | mri | t1c | lesion | met |
| Metastasis MRI T1 | mri | t1 | lesion | met |
| Normal CT Bone | ct | ctbo | normal | normal |
| Normal CT Brain | ct | ctbr | normal | normal |
| Normal MRI DWI | mri | dwi | normal | normal |
| Normal MRI Flair | mri | flair | normal | normal |
| Normal MRI T1 | mri | t1 | normal | normal |
| Normal MRI T1C+ | mri | t1c | normal | normal |
| Normal MRI T2 | mri | t2 | normal | normal |
| Schizophrenia MRI DWI | mri | dwi | diffuse | schiz |
| Schizophrenia MRI T1 | mri | t1 | diffuse | schiz |
| Glioma MRI T1C+ | mri | t1c | lesion | tumor |
| Meningioma MRI T1C+ | mri | t1c | lesion | tumor |
| Neurocitoma MRI T1C+ | mri | t1c | lesion | tumor |
| Other Lesions MRI T1C+ | mri | t1c | lesion | lesion |
| Schwannoma MRI T1C+ | mri | t1c | lesion | tumor |
| Glioma MRI T1 | mri | t1 | lesion | tumor |
| Meningioma MRI T1 | mri | t1 | lesion | tumor |
| Neurocitoma MRI T1 | mri | t1 | lesion | tumor |
| Other Lesions MRI T1 | mri | t1 | lesion | tumor |
| Schwannoma MRI T1 | mri | t1 | lesion | tumor |
| Glioma MRI T2 | mri | t2 | lesion | tumor |
| Meningioma MRI T2 | mri | t2 | lesion | tumor |
| Neurocitoma MRI T2 | mri | t2 | lesion | tumor |
| Other Lesions MRI T2 | mri | t2 | lesion | lesion |
| Schwannoma MRI T2 | mri | t2 | lesion | tumor |

† T1 MRI and T1 Contrast Enhanced MRI were designated a 0.5-unit difference. T2 MRI, DWI MRI, and Flair MRI were designated a 0.5-unit difference. All other difference between a class value was designated as a one-unit difference.

## II. Average Class Embedding Space Distance Heat Map

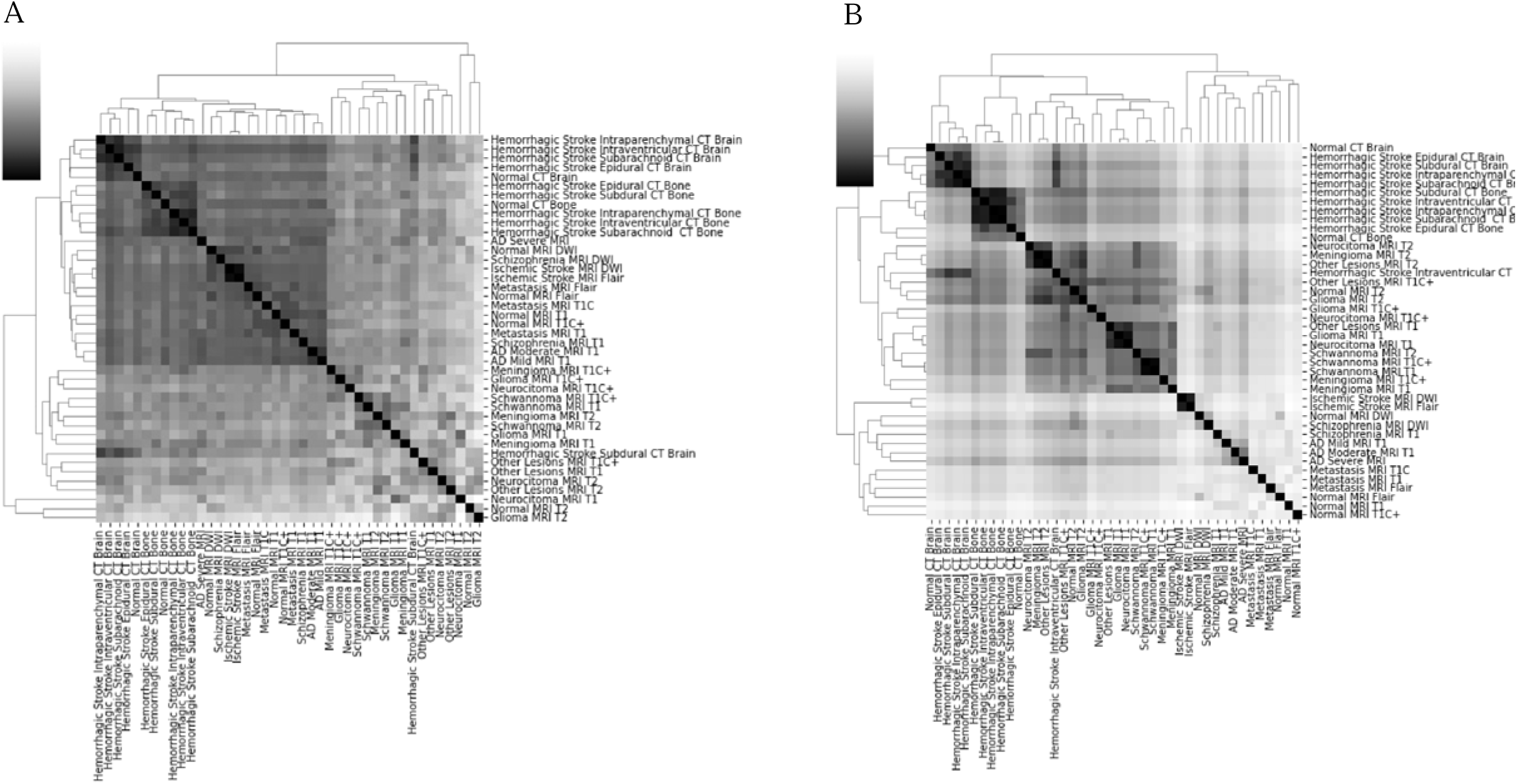
The figure illustrates the hierarchical clustering dendrogram of the average class embedding space of the Euclidean ResNet 18 (A) and the Euclidean-Lorentz ResNet 18 (B) in the Multi-Modality Neuroimaging (MMN) Dataset. The dendrogram is accompanies by a heatmap where the darker voxels represent a closer distance between the respective classes.

## III. Hierarchical Dendrograms in the Other Disease Datasets

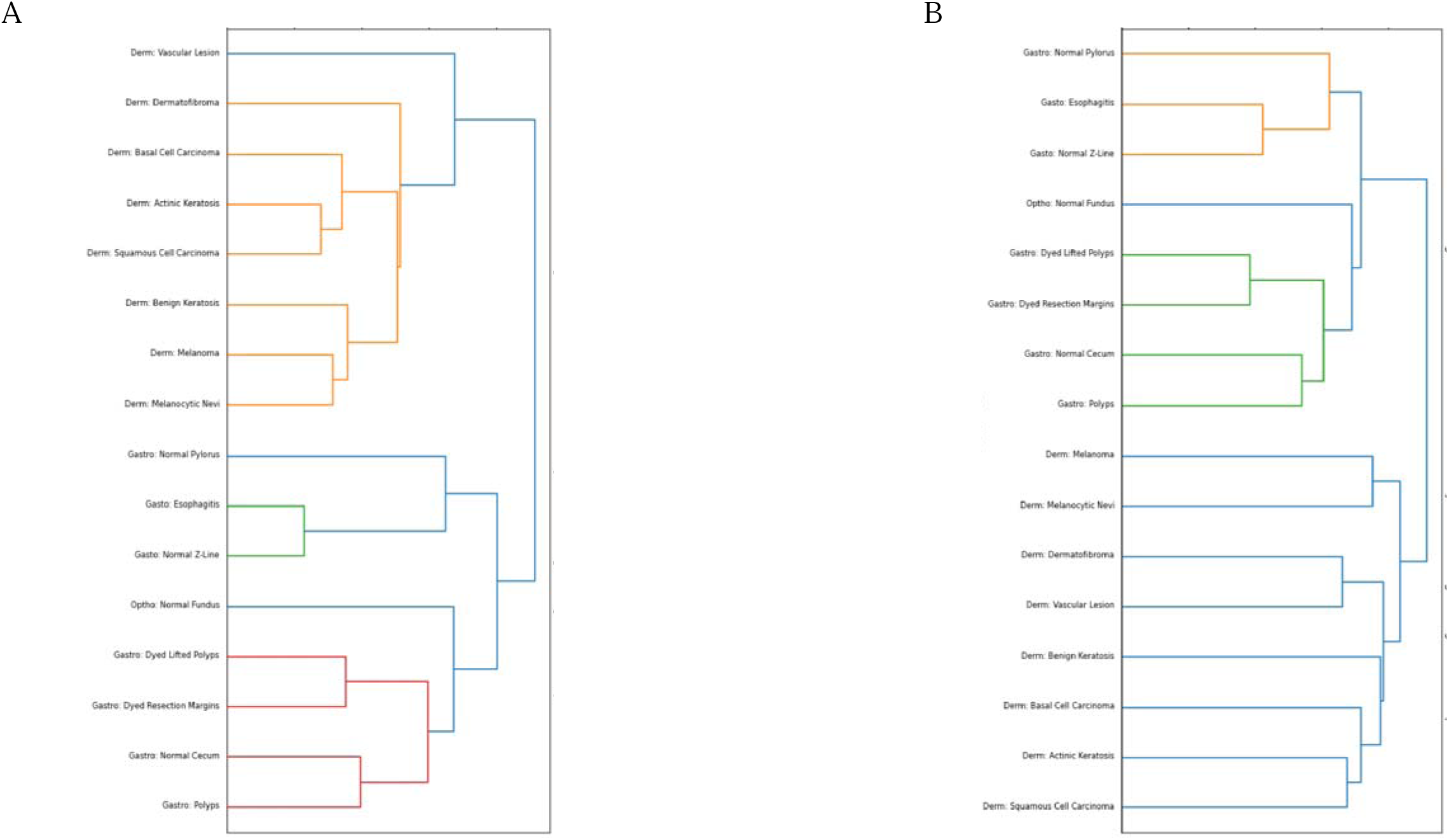
The figure illustrates the hierarchical clustering dendrogram of the average class embedding space of the Euclidean ResNet 18 (A) and the Euclidean-Lorentz ResNet 18 (B) in the Miniature Multi-Disease (MMD) Dataset.

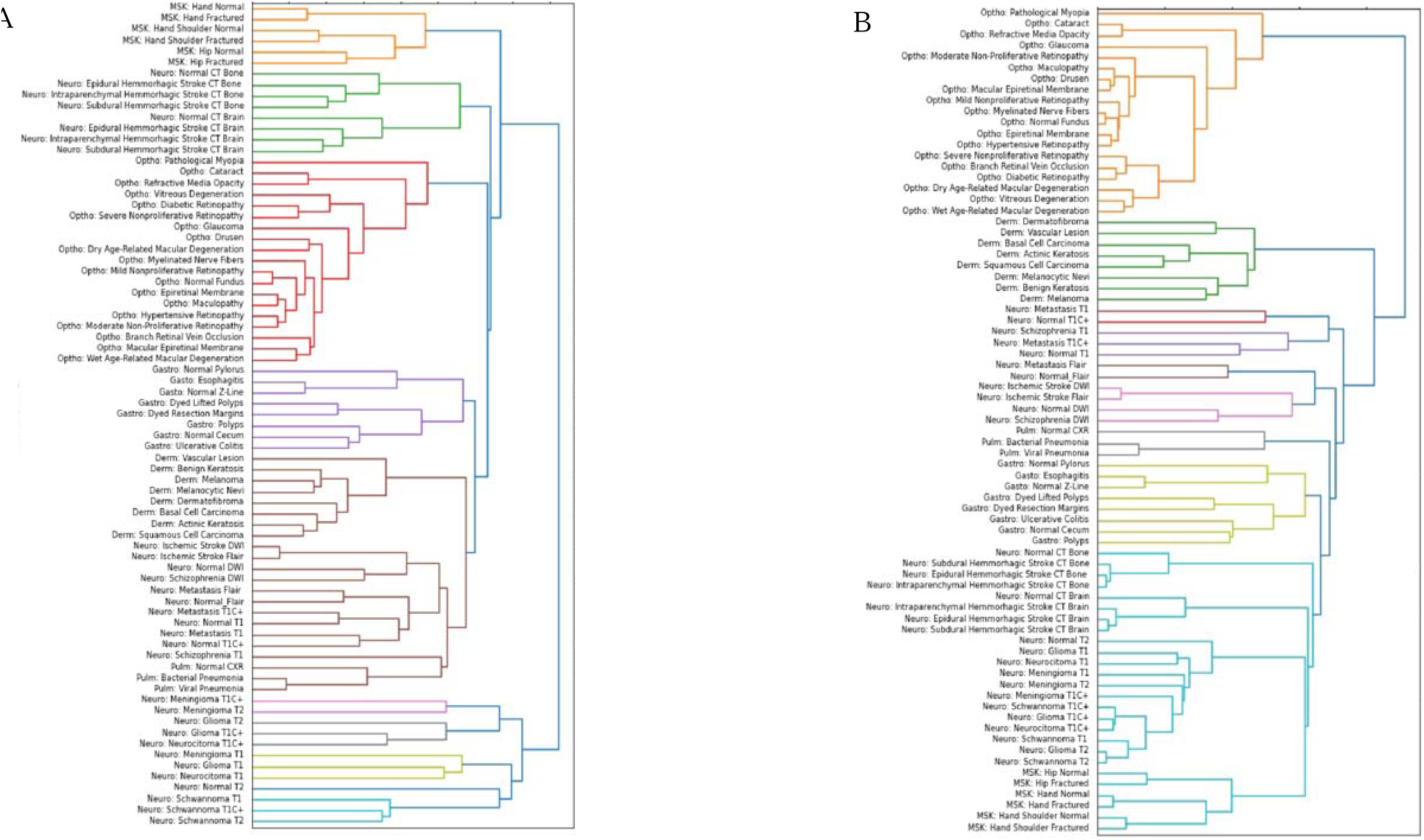
The figure illustrates the hierarchical clustering dendrogram of the average class embedding space of the Euclidean ResNet 18 (A) and the Euclidean-Lorentz ResNet 18 (B) in the Multi-Disease (MD) Dataset.

